# Longitudinal Tracking of Chromosomal Instability Informs Timely Intervention in the Gastric Precancerous Cascade

**DOI:** 10.1101/2025.11.09.25339830

**Authors:** Shao-wei Li, Xin-yu Fu, Chan Dai, Hao Liu, Wei-xia Wu, Jia-xiu Ying, Jin-bang Peng, Xian-bin Zhou, Bi-li He, Qin Huang, Min Zhu, Liang-min Zhang, Yu Zhang, Bin-bin Gu, Ling-ling Yan, Jia-cheng Li, Ren-quan Luo, Sai-qin He, Li-na Fang, Yan-di Lu, Ya-qi Song, Shi-wen Xu, Shen-ping Tang, Yu-han Lu, Shan-jing Xu, Xia Chen, Ya-hong Chen, Li-ping Ye, Mei-fu Gan, Xin-li Mao

**Affiliations:** Department of Gastroenterology, Taizhou Hospital of Zhejiang Province affiliated to Wenzhou Medical University, Linhai, Zhejiang, 317000, China; Zhejiang Provincial Clinical Research Center for Digestive Diseases, Linhai, Zhejiang, 317000, China; Institute of Digestive Disease, Taizhou Hospital of Zhejiang Province affiliated with Wenzhou Medical University, Linhai, Zhejiang, 317000, China; Key Laboratory of Minimally Invasive Techniques & Rapid Rehabilitation of Digestive System Tumor of Zhejiang Province, Taizhou Hospital Affiliated to Wenzhou Medical University, Linhai, Zhejiang, 317000, China; Dalian Medical University, Dalian, China; State Key Laboratory of Genetic Engineering, Fudan Microbiome Center, Human Phenome Institute, Fudan University, Shanghai, China; Department of Surgery, Taizhou Hospital of Zhejiang Province Affiliated to Wenzhou Medical University, Linhai, Zhejiang, 317000, China; Department of Emergency Medicine, The Second People’s Hospital of Linhai City, Linhai, 317016, Zhejiang, China; Taizhou Hospital of Zhejiang Province, Shaoxing University, Shaoxing, Zhejiang, China; Taizhou Hospital of Zhejiang Province Affiliated to Wenzhou Medical University, Linhai, Zhejiang 317000, China; Department of Pathology, Taizhou Hospital of Zhejiang Province affiliated to Wenzhou Medical University, Linhai, Zhejiang, 317000, China; Taizhou Enze Medical Center(Group) (Taizhou University Affiliated Enze Medical Center), School of Medicine, Taizhou University, Linhai, Zhejiang, 317000, China; The Johns Hopkins University-Krieger School of Arts & Sciences, Baltimore, Maryland 21218, United States; Department of Gastroenterology, Wenling First People’s Hospital, Taizhou, Zhejiang 317500, China; Health Management Center, Taizhou Hospital of Zhejiang Province Affiliated to Wenzhou Medical University, Linhai, Zhejiang, 317000, China

**Keywords:** Chromosomal instability, Endoscopic submucosal dissection, Gastric cancer

## Abstract

**Background:** A major unmet need in gastric cancer prevention is the lack of biomarkers to predict precancerous lesion progression, and the potential of chromosomal instability (CIN) for this role, particularly regarding early cancer and recurrence, remains unclear.

**Objective:** To assess the predictive role of CIN in lesion progression and postoperative recurrence following endoscopic submucosal dissection (ESD).

**Design:** We enrolled 106 patients from Taizhou Hospital Affiliated to Wenzhou Medical University (2011–2025) and collected 1,045 temporally continuous pathological samples.

Copy number variations were profiled using low-coverage whole-genome sequencing (LC-WGS), and CIN scores were calculated. Associations between CIN and clinical outcomes were analyzed using survival analysis and ROC curves.

**Results:** CIN was detectable up to two years before gastric cancer diagnosis, with positivity rates increasing alongside lesion severity: 25.5% in intestinal metaplasia, 39.7% in low-grade dysplasia, and 83.0% in gastric cancer. CIN positivity independently predicted lesion progression (hazard ratio [HR] = 2.55, 95% CI: 1.19–5.47, p = 0.016), with the greatest risk for progression to carcinoma (HR = 16.61, p < 0.001). The predictive AUC was 0.81, improving to 0.88 when combined with age. Among 84 patients who underwent ESD, CIN positivity significantly increased recurrence risk (HR = 19.57, 95% CI: 2.59 – 147.61, p = 0.004; AUC = 0.80). Chromosomal arms 7p/q, 8p/q, and 20p/q showed high CIN frequencies, with MYC being the most frequently mutated oncogene.

**Conclusion:** CIN represents a reliable biomarker for early prediction of lesion progression and postoperative recurrence, enabling proactive surveillance and precision management of gastric cancer.

**Single Sentence Summary:** Chromosomal instability (CIN) acts as a reliable biomarker for predicting the progression of gastric precancerous lesions and postoperative recurrence.

## INTRODUCTION

Gastric cancer remains a leading cause of cancer-related mortality worldwide, with marked geographical variations in incidence and prognosis [1, 2]. Early detection and accurate diagnosis are crucial for improving outcomes, as timely intervention significantly enhances survival rates. Gastric carcinogenesis typically follows a multistep progression, beginning with chronic gastritis and advancing through atrophic gastritis, intestinal metaplasia, low-grade dysplasia, and high-grade dysplasia, ultimately leading to invasive carcinoma [3, 4]. The transition from intestinal metaplasia and low-grade dysplasia to early gastric cancer (EGC) is particularly critical, as intervention at this stage can effectively halt disease progression [3–5]. However, the current diagnosis and management of precancerous lesions, such as intestinal metaplasia, still rely heavily on histomorphological assessment, which is limited by subjectivity and interobserver variability. These constraints hinder consistent and accurate evaluation of malignant potential. Thus, there is an urgent need for predictive molecular biomarkers that can objectively assess the risk of progression, enabling more precise risk stratification and facilitating a shift from passive surveillance to proactive monitoring and intervention.

Chromosomal instability (CIN), a common genomic alteration in cancer cells, is characterized by the accumulation of structural and numerical chromosomal abnormalities and is a major driver of tumor initiation and progression [6, 7]. In gastric cancer, CIN promotes malignant transformation by increasing genetic diversity and accelerating clonal evolution [6]. Recent studies suggest that CIN may serve as a biomarker for identifying high-risk precancerous lesions [8–10]. For example, genomic analyses have shown that chromosomal aberrations detected in precancerous gastric lesions are associated with progression to EGC, underscoring the predictive value of CIN in malignant transformation [11, 12]. Incorporating CIN assessment into clinical practice could enhance the risk stratification of precancerous lesions and guide treatment decisions, thereby improving early prevention and control strategies for gastric cancer. Advances in genomic technologies, such as comparative genomic hybridization and high-throughput sequencing, have made CIN detection feasible, yet standardization of analytical methods and validation across diverse populations remain necessary.

This study aimed to systematically investigate the role of CIN in the early stages of gastric carcinogenesis, focusing on its predictive value for identifying precancerous lesions at high risk of progression. By analyzing a longitudinal cohort spanning intestinal metaplasia to gastric cancer, we sought to clarify the association between CIN and malignant transformation and to establish CIN as a reliable early diagnostic biomarker. Ultimately, this work aims to contribute to the development of precise diagnostic tools for high-risk populations and to provide a foundation for improving clinical outcomes.

## MATERIALS AND METHODS

### Sample preparation

All samples were obtained with approval from the Ethics Committee of Taizhou Hospital Affiliated to Wenzhou Medical University (Approval No. K20201205). Written informed consent was obtained from all patients for prospective tissue collection. Hematoxylin and eosin (H&E)-stained sections were independently reviewed by at least three expert pathologists.

This study included a longitudinal cohort of patients enrolled between 2011 and 2025. The inclusion criteria were: (i) completion of two or more endoscopic examinations at the study center; (ii) confirmation of intestinal metaplasia in at least one pathological examination; (iii) availability of complete clinical and pathological information; and (iv) integrity of formalin-fixed paraffin-embedded (FFPE) tissue blocks meeting quality standards for analysis.

Based on pathological characteristics, the cohort was further divided into progression and recurrence groups. The progression group included patients with: (i) an initial pathological sample not obtained during surgery and not intended for preoperative evaluation, and (ii) subsequent follow-up (same or different anatomical site) showing progression to a high-risk lesion. The recurrence group included patients with: (i) a pathological sample obtained via endoscopic submucosal dissection (ESD) and (ii) postoperative endoscopic follow-up with subsequent pathological sampling.

High-risk lesions were defined as those exhibiting malignant features or malignant potential, including low-grade intraepithelial neoplasia (LGIN), high-grade intraepithelial neoplasia (HGIN), and EGC [13].

Disease progression was defined as the development of a high-risk lesion at the same or a different anatomical site during follow-up. Recurrence was defined as the reappearance of a high-risk lesion at the same or a different site after ESD.

### DNA extraction

Genomic DNA was extracted from FFPE tissue sections using the QIAamp DNA Mini Kit (Qiagen, Germany) according to the manufacturer’s instructions to ensure consistency across samples. DNA concentration, purity, and integrity were assessed using a NanoView spectrophotometer (GE Healthcare). DNA quality was further verified by 0.8% agarose gel electrophoresis to confirm suitability for downstream applications.

### Low-coverage whole-genome sequencing

Low-coverage whole-genome sequencing (LC-WGS) was performed using the Kapa Hyper Prep Kit (Roche, Basel, Switzerland) with IDT-specific adapters. DNA input ranged from 50 to 800 ng. A total of 22 libraries were pooled and sequenced using a paired-end 150 bp strategy on an Illumina HiSeq X10 platform. Copy number variations (CNVs) were inferred using the “Ultra-Sensitive Aneuploidy Detection” pipeline. For quality control, samples were excluded if the median absolute deviation (MAD) of copy ratios between adjacent genomic bins exceeded 0.38.

### Copy number analysis and CIN scoring

CNVs and breakpoints were identified using the circular binary segmentation (CBS) algorithm implemented in the R package DNACopy. Each sample generated at least 10 million paired-end reads aligned to the hg19 reference genome. Average coverage was calculated in 200 kb bins using *Samtools mpileup*, normalized, and converted into Z-scores. Regions with Z-scores > 3 were defined as amplified, and those with Z-scores < –3 as deleted. CIN was assessed based on these alterations.

### Statistical analysis

The Mann–Whitney U test was used for comparisons between two independent groups, and the Kruskal–Wallis test for comparisons across more than two groups. Fisher’s exact test was applied to compare mutation frequencies and arm-level copy number alteration (CNA) frequencies between groups. The Benjamini–Hochberg method was used to control the false discovery rate, with a q-value < 0.05 considered statistically significant. Additional details are provided in the Supplementary Materials and Methods (Supplementary Material 1).

### Patient and Public Involvement

Patients or members of the public were not involved in the design, conduct, reporting, or dissemination plans of this research.

## RESULTS

### Sample characteristics

A total of 1,199 longitudinally collected pathological specimens from 107 patients were included in this study. After excluding samples that failed quality control or sequencing, 1,045 samples from 106 patients were retained for the final analysis (Figure 1). The mean number of samples per patient was 9.8, with a median of nine samples per patient. CIN was detected in 280 samples (26.8%) classified as CIN-positive. These samples exhibited a median of five aberrant chromosomal arms with deletions and 4,365 mutated genes. Among the pathological subtypes, CIN was identified in 39 of 47 cancer samples (83.0%), 62 of 156 dysplasia samples (39.7%), and 128 of 501 intestinal metaplasia samples (25.5%). The detailed characteristics are summarized in Table 1. The timeline of all samples is shown in Figures S1–S4.

**Figure 1.**
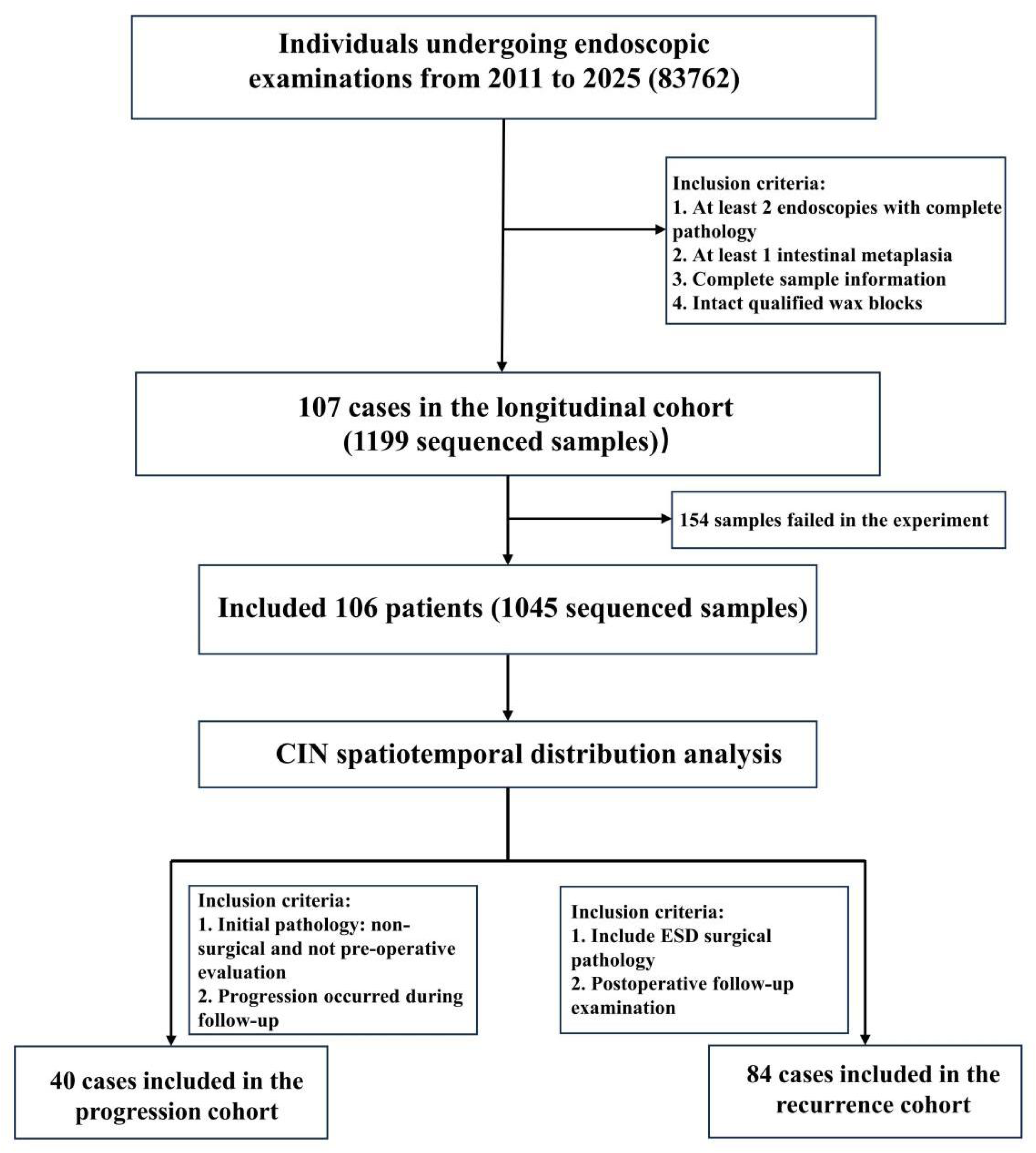
Study flowchart.

**Table 1.** Characteristic Distribution of Gastric-Related Pathological Samples Under Different Chromosomal Instability (CIN) Statuses.

| Variable | Overall<br>(n=1,045) | CIN (-)<br>(n=765) | CIN (+)<br>(n=280) | *P* |
| --- | --- | --- | --- | --- |
| Degree of lesion, n (%) |  |  |  | <0.001 |
| CSG | 299 (28.61%) | 259 (33.86%) | 40 (14.29%) |  |
| IM | 501 (47.94%) | 373 (48.76%) | 128 (45.71%) |  |
| GIN | 156 (14.93%) | 94 (12.29%) | 62 (22.14%) |  |
| GC | 47 (4.50%) | 8 (1.04%) | 39 (13.93%) |  |
| Other | 42 (4.02%) | 31 (4.05%) | 11 (3.93%) |  |
| Location of lesion, n (%) |  |  |  | <0.001 |
| AR | 170 (16.27%) | 119 (15.56%) | 51 (18.21%) |  |
| CA | 510 (48.80%) | 412 (53.86%) | 98 (35.00%) |  |
| CB | 183 (17.51%) | 122 (15.95%) | 61 (21.79%) |  |
| CD | 167 (15.98%) | 103 (13.46%) | 64 (22.86%) |  |
| SF | 13 (1.24%) | 9 (1.18%) | 4 (1.43%) |  |
| AS | 2 (0.02%) | 0 (0%) | 2 (0.07%) |  |
| Chromosome Arm Number, Median (Q1, Q3) | 0.0 (0.0, 1.0) | 0.0 (0.0, 0.0) | 5.0 (2.5, 9.0) | <0.001 |
| HP infection level |  |  |  | 0.823 |
| + | 68 (6.51%) | 47 (6.14%) | 21 (2.68%) |  |
| ++ | 34 (3.25%) | 24 (3.14%) | 10 (3.57%) |  |
| +++ | 13 (1.24%) | 10 (1.31%) | 3 (1.07%) |  |
| Negative | 930 (89.00%) | 684 (89.41%) | 246 (87.86%) |  |
| Number of gene, Median (Q1, Q3) | 0 (0, 873) | 0 (0, 0) | 4,365 (2,183, 7,857) | <0.001 |
| Chr 1 | 41 (3.92%) | 0 (0%) | 41 (14.64%) | <0.001 |
| Chr 2 | 55 (5.26%) | 0 (0%) | 55 (19.64%) | <0.001 |
| Chr 3 | 46 (4.40%) | 0 (0%) | 46 (16.07%) | <0.001 |
| Chr 4 | 34 (3.25%) | 0 (0%) | 34 (12.14%) | <0.001 |
| Chr 5 | 53 (5.07%) | 0 (0%) | 53 (18.93%) | <0.001 |
| Chr 6 | 51 (4.88%) | 0 (0%) | 51 (4.88%) | <0.001 |
| Chr 7 | 98 (9.38%) | 0 (0%) | 98 (35%) | <0.001 |
| Chr 8 | 123 (11.77%) | 0 (0%) | 123 (43.93%) | <0.001 |
| Chr 9 | 57 (5.45%) | 0 (0%) | 57 (20.38%) | <0.001 |
| Chr 10 | 49 (4.59%) | 0 (0%) | 49 (17.50%) | <0.001 |
| Chr 11 | 23 (2.20%) | 0 (0%) | 23 (8.21%) | <0.001 |
| Chr 12 | 51 (4.88%) | 0 (0%) | 51 (18.21%) | <0.001 |
| Chr 13 | 110 (10.53%) | 0 (0%) | 110 (39.29%) | <0.001 |
| Chr 14 | 52 (4.97%) | 0 (0%) | 52 (18.57%) | <0.001 |
| Chr 15 | 29 (2.78%) | 0 (0%) | 29 (10.36%) | <0.001 |
| Chr 16 | 36 (3.44%) | 0 (0%) | 36 (12.86%) | <0.001 |
| Chr 17 | 47 (4.5%) | 0 (0%) | 47 (17.0%) | <0.001 |
| Chr 18 | 47 (4.50%) | 0 (0%) | 47 (16.79%) | <0.001 |
| Chr 29 | 3 (0.29%) | 0 (0%) | 3 (1.07%) | <0.001 |
| Chr 20 | 113 (10.81%) | 0 (0%) | 113 (40.36%) | <0.001 |
| Chr 21 | 14 (1.34%) | 0 (0%) | 14 (5.00%) | <0.001 |
| Chr 22 | 32 (3.06%) | 0 (0%) | 32 (11.43%) | <0.001 |

### Patient characteristics

The analyzed cohort comprised 106 patients (69 males and 37 females) with a median age of 65 years. Comorbidities and risk factors included hypertension (n = 31), diabetes (n = 10), history of *Helicobacter pylori* infection (n = 68), smoking (n = 40), alcohol consumption (n = 19), and illiteracy (n = 29). During follow-up, 84 patients had at least one CIN-positive sample. Notably, all six patients with a family history of gastric cancer were positive for CIN. Moreover, the median age of CIN-positive patients was significantly higher than that of CIN-negative patients (p = 0.003). Further details are presented in Table 2.

**Table 2.** Clinical Baseline Characteristics of Patients with Gastric Diseases Under Different Chromosomal Instability (CIN) Statuses.

| Variable | Overall<br>(n=106) | CIN (-)<br>(n=22) | CIN (+)<br>(n=84) | *P* |
| --- | --- | --- | --- | --- |
| Age, median (Q1, Q3), years | 69 (65, 74) | 66 (60, 68) | 72 (66, 76) | 0.003 |
| Sex, n (%) |  |  |  | 0.399 |
| Male | 69 (65.09%) | 16 (72.73%) | 53 (63.10%) |  |
| Female | 37 (34.91%) | 6 (27.27%) | 31 (36.90%) |  |
| Family history of gastric cancer, n (%) | 6 (5.66%) | 0 (0%) | 6 (7.14%) | 0.341 |
| Hypertension, n(%) | 31 (29.25%) | 5 (22.73%) | 26 (30.95%) | 0.450 |
| Diabetes, n(%) | 10 (9.43%) | 3 (13.63%) | 7 (8.33%) | 0.430 |
| History of HP infection, n(%) | 68 (64.15%) | 11 (50.00%) | 57 (67.86%) | 0.120 |
| Smoking history, n(%) | 40 (37.74%) | 10 (45.45%) | 30 (35.71%) | 0.401 |
| Alcohol use history, n(%) | 19 (17.92%) | 6 (27.27%) | 13 (15.48%) | 0.219 |
| Education Background, n(%) |  |  |  | 0.584 |
| Illiterate | 29 (27.36%) | 5 (22.73%) | 24 (28.57%) |  |
| Primary Education or Higher | 77 (72.64%) | 17 (77.27%) | 60 (71.43%) |  |
| Occupation, n(%) |  |  |  | 0.124 |
| Retired Status | 13 (12.26%) | 2 (9.09%) | 11 (13.10%) |  |
| Farmer | 79 (74.53%) | 14 (63.64%) | 65 (77.38%) |  |
| Worker | 14 (13.21%) | 6 (27.27%) | 8 (9.52%) |  |
| BMI, Median (Q1, Q3) | 23.23 (20.87, 24.81) | 24.55 (21.63, 25.31) | 23.11 (20.56, 24.57) | 0.157 |

### Temporal and spatial specificity of CIN

Sequencing data revealed a significant correlation between the extent of CIN and the progression of gastric lesions. As the lesions advanced from benign to malignant stages, the degree of CIN demonstrated a stepwise increase(CSG 13.38%, IM 25.55%,GIN 39.74%, GC 83.00%), with gastric cancer showing the highest CIN positivity rate and mutational burden (Figure 2B). Notably, CIN was already detectable at the intestinal metaplasia stage, indicating that its emergence precedes gastric cancer development and suggesting its important role in the early phases of gastric carcinogenesis.

**Figure 2.**
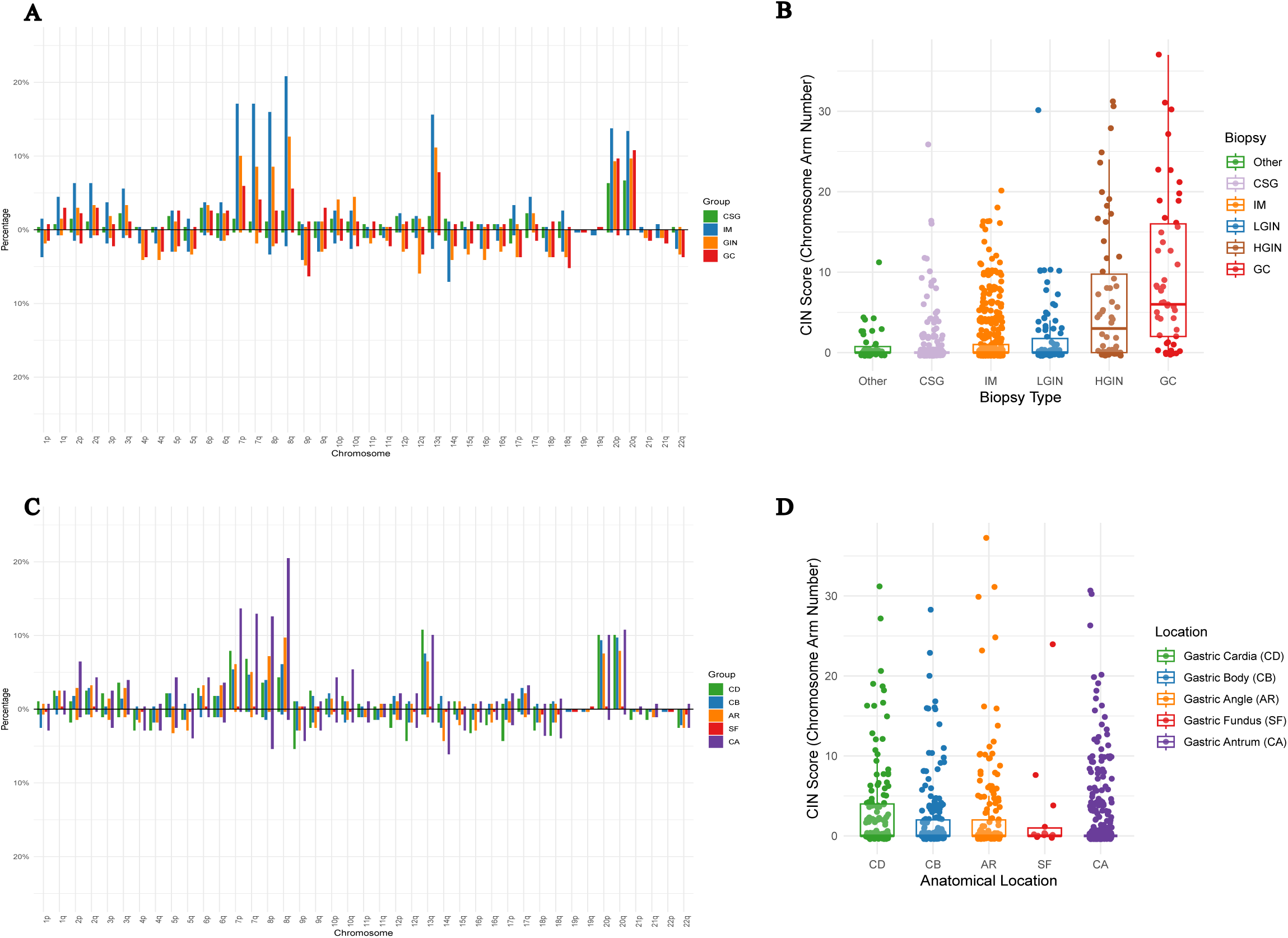
Spatiotemporal distribution of chromosome instability (CIN). A. Distribution of chromosomal mutations across pathological stages; B. Degree of chromosomal mutations in lesions; C. Distribution of mutations across different chromosomal regions; D. Degree of chromosomal mutations across anatomical sites.

We focused on four key pathological stages—gastritis, intestinal metaplasia, dysplasia, and gastric cancer—to investigate the distribution patterns of chromosomal alterations. CIN events at the intestinal metaplasia stage displayed broad genomic coverage, with particularly high frequencies of aberrations in chromosomal arms 7p, 7q, 8p, 8q, 13q, 20p, and 20q (Figure 2A). These specific chromosomal abnormalities may serve as potential biomarkers for the early detection of gastric cancer and precancerous conditions and could also represent targetable sites for future therapeutic interventions.

Spatial analysis of CIN revealed notable specificity across different gastric regions. The highest CIN positivity rate was observed in the cardia (38.3%), which may be attributed to its unique anatomical position as a transition zone between the esophageal squamous epithelium and the gastric glandular epithelium—a histological context that potentially predisposes patients to genomic instability. In contrast, the antrum showed a lower CIN positivity rate (19.2%) compared with the gastric angle (30.0%), body (33.3%), or fundus (30.8%) (Figure 2D). However, within CIN-positive cases in the antrum, a higher proportion of mutations involved chromosomes 7 and 8 (Figure 2C). Overall, the spatial distribution of CIN correlated strongly with both regional susceptibility and the severity of the mutational burden.

We analyzed the mutated genes on the chromosome (Figure S5). The distribution pattern of mutated genes across disease severity stages was consistent with that of CIN, showing a stepwise increase in mutational burden. The median numbers of mutated genes were 4,387 in chronic gastritis, 4,638 in intestinal metaplasia, 7,083 in dysplasia, and 9,536 in gastric cancer (Figure 3A). The gastric fundus exhibited the highest median number of mutated genes (7,202), followed by the gastric angle (6,539), indicating significant differences in genomic instability among the anatomical subsites of the stomach (Figure 3B).

**Figure 3.**
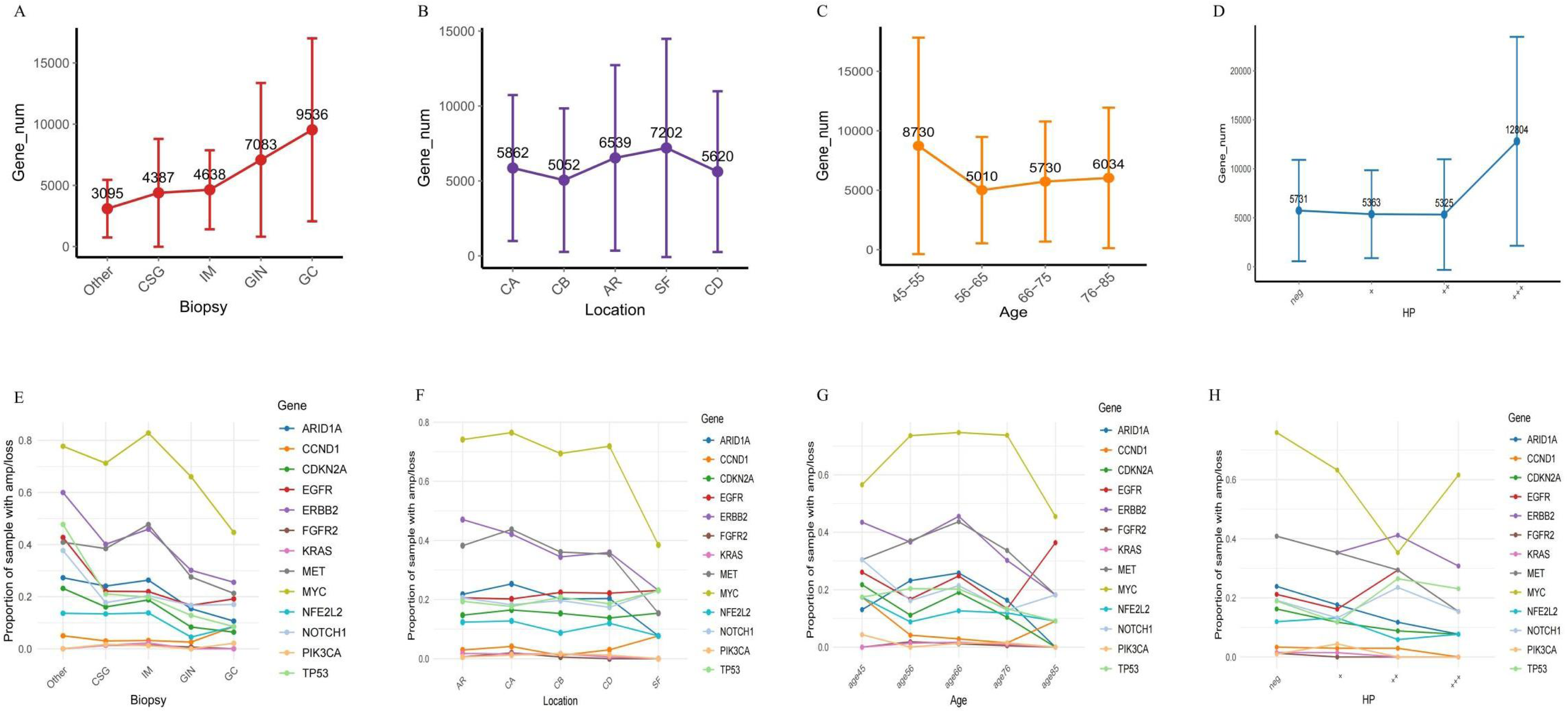
Distribution of mutated genes. A. Mutated genes across pathological stages; B. Mutated genes across anatomical regions; C. Mutated genes across age groups; D. Mutated genes according to *Helicobacter pylori* infection status; E. Oncogenes across pathological stages; F. Oncogenes across anatomical regions; G. Oncogenes across age groups; H. Oncogenes according to *H. pylori* infection status.

Age-stratified analysis revealed a peak in the median number of mutated genes (8,730) in the 45–55 year age group, suggesting that this period may represent a critical window for gastric lesion progression (Figure 3C). Furthermore, stratification by *H. pylori* infection status demonstrated that the heavily infected group (+++) had a significantly higher mutational burden than other groups (Figure 3D), consistent with the known role of persistent *H. pylori* infection in promoting genomic damage in the gastric mucosa.

In the specificity analysis of relevant oncogenes, MYC consistently exhibited the highest mutation frequency across all disease stages, anatomical subsites, age subgroups, and *H. pylori* infection strata (Figure 3E-H). Its ubiquitous mutational prevalence was observed across different anatomical regions and remained stably high regardless of age or infection intensity. Notably, the MYC mutation frequency was particularly elevated in intestinal metaplasia, reinforcing the hypothesis that highly proliferative metaplastic lesions may possess greater malignant potential. These findings indicate that aberrant activation of the MYC signaling pathway may represent a central molecular event in gastric lesion progression. Thus, targeting MYC and its downstream effectors could offer novel, precise therapeutic strategies for reversing precancerous gastric conditions and treating gastric cancer.

### CIN as a predictive biomarker for progression

Based on predefined criteria for disease progression, 40 patients were included in the progression cohort. Among them, 29 (72.50%) exhibited disease progression with a CIN positivity rate of 62.09%. Specifically, progression to gastric cancer occurred in five cases (CIN positivity: 80.00%), low-grade dysplasia in 12 cases (CIN positivity: 38.46%), and high-grade dysplasia in 12 cases (CIN positivity: 75.00%).

CIN positivity was identified as an independent risk factor for lesion progression (hazard ratio [HR] = 2.55, 95% CI: 1.19–5.47, p = 0.016), with a median progression time of 24.27 months (Figure 4A). Stratified analysis revealed particularly high risks for progression to gastric cancer (HR = 16.61, 95% CI: 1.83–150.60, p < 0.001; median progression time: 15.88 months) (Figure S6A), to high-grade intraepithelial neoplasia (HR = 4.23, 95% CI: 1.11–16.06, p = 0.022; median progression time: 34.03 months) (Figure S6B), and to low-grade intraepithelial neoplasia (HR = 5.69, 95% CI: 1.61–20.11, p = 0.003; median progression time: 19.36 months) (Figure S6C).

**Figure 4.**
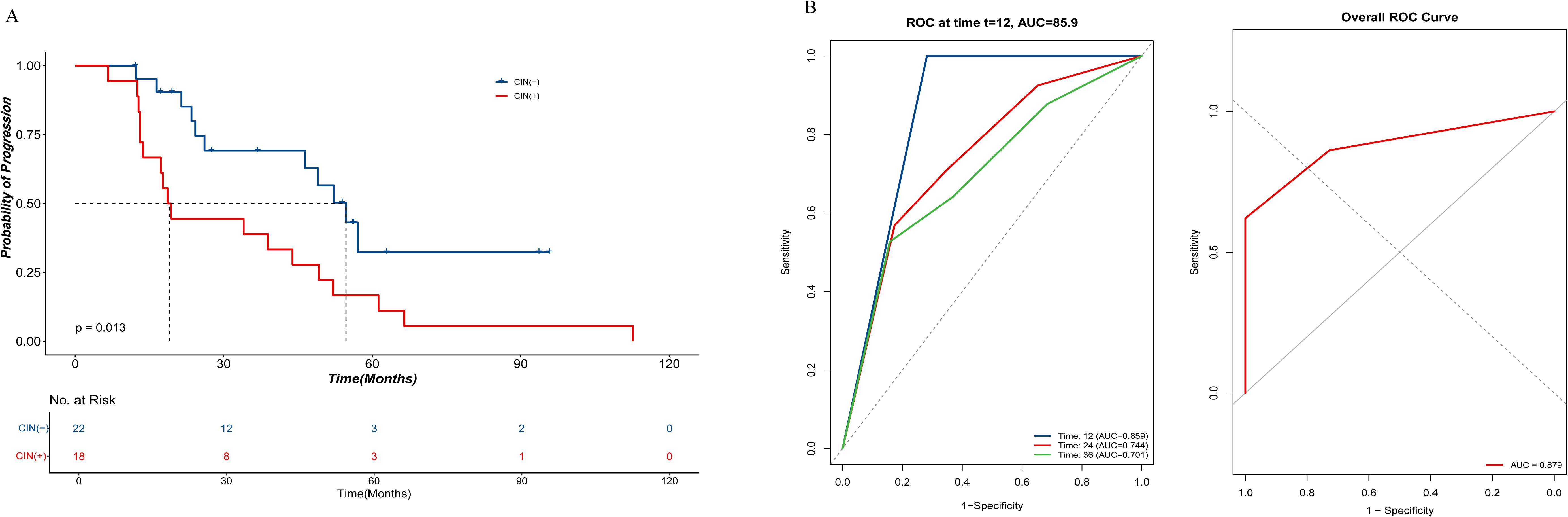
Kaplan–Meier and ROC curves for the progression cohort. A. Kaplan–Meier curve of progression risk in CIN-positive patients; B. ROC curve for predicting progression using CIN.

The predictive performance of CIN for progression was evaluated using time-dependent ROC analysis, yielding an AUC of 0.81. The AUCs for predicting progression within the first, second, and third years were 0.78, 0.68, and 0.64, respectively (Figure S7A). These results indicate that CIN serves not only as an early warning marker for gastric cancer development but also reflects persistent instability closely associated with disease progression. This provides a critical rationale for early intervention: by monitoring CIN in precancerous states such as intestinal metaplasia, high-risk individuals can be identified early, shifting the intervention window to the preclinical phase.

In analyses of other potential progression-related factors, older age showed a trend toward being a risk factor (HR = 2.01, 95% CI: 0.92–4.38, p = 0.081) (Figure S6D). When adjusted for age in the multivariate model, the predictive accuracy of CIN improved, with the AUC increasing to 0.88 (Figure 4B). There were no significant differences in age or *H. pylori* infection. Although *H. pylori* infection is a known risk factor for cancer, it did not show a significant association with cancer progression in this cohort (Table S1). This may be due to limitations in accurately assessing infection status during long-term follow-up, which could affect the reliability of its correlation with disease progression.

A noteworthy subgroup consisted of 12 patients exhibiting postoperative pathological upstaging, predominantly carcinoma (11 cases, 91.67%), among whom the CIN positivity rate was 90.91%. Pathological upgrading was observed from high-grade dysplasia to carcinoma (six cases), low-grade dysplasia to carcinoma (two cases), and intestinal metaplasia to carcinoma (three cases). These findings highlight the limitations of relying solely on histopathological evaluations, which may be influenced by sampling variability during endoscopy. CIN detection may serve as a corrective tool to mitigate such diagnostic uncertainties.

### CIN as a risk factor for recurrence

Based on longitudinal cohort data, persistent CIN was significantly associated with adverse clinical outcomes. Among the 84 patients included in the recurrence cohort, those with CIN-positive postoperative pathological samples had a markedly higher risk of high-risk lesion recurrence (HR = 19.57, 95% CI: 2.59–147.61, p = 0.004), with a median recurrence time of 28.27 months (Figure 5A). Thus, CIN status serves as an independent biological predictor of postoperative recurrence. The predictive performance, as measured by AUC, was 0.76. Time-dependent AUC values for recurrence prediction were 0.75, 0.76, and 0.76 for the first, second, and third years, respectively (Figure S7B).

**Figure 5.**
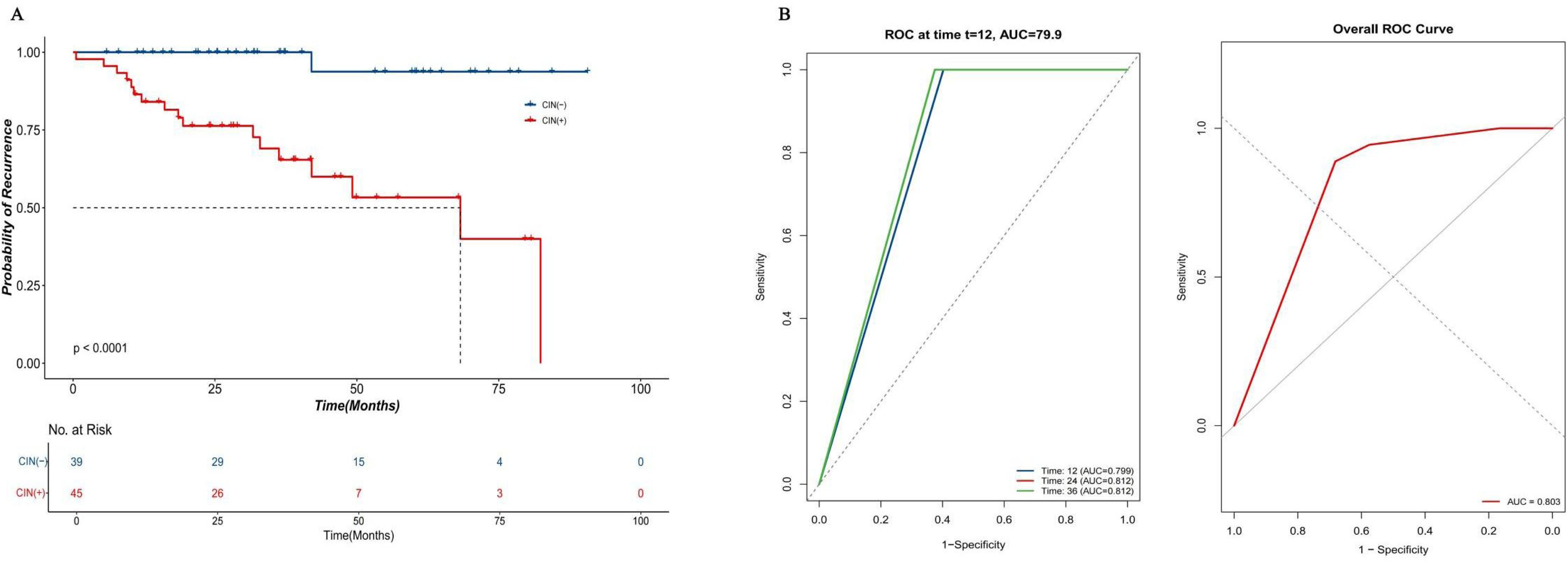
Kaplan–Meier and ROC curves for the recurrence cohort. A. Kaplan–Meier curve of recurrence risk in CIN-positive patients; B. ROC curve for predicting recurrence using CIN.

Age-stratified analysis indicated that patients aged > 65 years had a significantly higher risk of recurrence than younger individuals (p = 0.046) (Figure S6E). A combined model incorporating both CIN and age improved the predictive AUC to 0.80 (Figure 5B). In contrast, no significant association was observed between *H. pylori* infection status, sex, and recurrence (Table S1).

In the recurrence group, the CIN positivity rate was 94.44% (with one negative case likely attributable to sampling bias). Interestingly, a CIN positivity rate of 23.7% was also observed among non-recurrent patients, suggesting that CIN status may exhibit dynamic variability. Because this study relied on static sample assessments, it remains unclear whether CIN-positive cells can be cleared or repaired by the immune system. Therefore, persistent CIN, rather than transient positivity, may more accurately reflect high-risk progression. The 94.44% recurrence rate among CIN-positive patients underscores the clinical importance of long-term endoscopic surveillance and genomic monitoring for the early detection of recurrence and malignant transformation.

### Clonal evolution

Clonal evolution analysis was performed in patients with disease progression. These evolutionary patterns were largely consistent with the Correa cascade model, supporting its biological relevance in gastric carcinogenesis. However, not all trajectories strictly followed this model, indicating a degree of stochasticity in the process.

Mutation heatmaps revealed that chromosomes 7, 8, and 20 harbored the most frequently shared mutations during disease progression, highlighting their pivotal roles. These findings were validated in a cohort of patients who experienced recurrence. Recurrent genomic alterations on chromosomes 7, 8, and 20 may represent early events in gastric carcinogenesis. Thus, early detection and management of these alterations could provide an effective strategy for preventing both the development and recurrence of gastric cancer (Figures S8–S9).

## DISCUSSION

Early diagnosis and treatment of gastric cancer remain the most effective strategies for reducing disease burden [2]. A central scientific challenge lies in identifying key molecular events in gastric carcinogenesis and developing reliable methods for early detection and risk prediction. Accurate assessment of the progression risk from precancerous lesions to invasive carcinoma would enable proactive and individualized disease management. Based on a 15-year longitudinal endoscopic follow-up cohort, we identified a subpopulation with disease progression and performed whole-genome sequencing (WGS) of archived tissue samples. This approach established CIN as a key molecular marker in gastric cancer development and prognosis. Importantly, CIN was detectable at the precancerous stage of intestinal metaplasia (IM), suggesting its role as an early driver of malignant transformation.

The classical Correa cascade describes gastric mucosal carcinogenesis as a multistep process, with atrophy and IM as critical stages [3, 5]. Although recent studies have questioned the direct carcinogenic role of IM, our data reaffirm its importance. We detected widespread chromosomal alterations in IM samples, including high mutation frequencies on chromosomes 7, 8, 13, and 20, which closely resembled those in dysplasia and gastric cancer (GC). This suggests that IM clones with such alterations may represent the cellular origin of GC. While large-scale genomic studies have previously reported frequent alterations on chromosomes 8 and 13 in GC [14], our study confirmed these findings and identified recurrent alterations on chromosomes 7 and 20 as equally important, expanding the known genomic landscape of gastric carcinogenesis. Clonal evolutionary analysis further supported the Correa cascade in most cases, though exceptions were observed, underscoring the importance of early risk stratification in GC prevention.

In our cohort, CIN-positive patients had a 2.55-fold higher risk of lesion progression than CIN-negative individuals, with a median progression time of approximately 24 months. This indicates that high-risk populations can be identified at least two years before pathological progression. Although the annual malignant transformation rate of IM is relatively low (0.09–1.7%), its cumulative risk remains clinically relevant, and reliable biological tools for progression risk assessment are lacking [3, 15]. Our findings show that CIN is not only detectable at the IM stage but also increases with pathological advancement, indicating synchronization between CIN burden and disease severity. ROC analysis revealed that combining CIN with age achieved an AUC of 0.879 for predicting progression risk, supporting its potential as a predictive biomarker.

Recent liquid biopsy technologies, such as assays based on exosomal lncRNAs or methylation markers in peripheral blood, show promise for early GC detection [16–19]. However, endoscopic biopsy and histopathology remain the diagnostic gold standard. Moreover, most existing biomarkers detect early cancer rather than predict precancerous lesion progression [19, 20]. Since prognosis strongly depends on timing, early identification of high-risk lesions is crucial. Endoscopic resection is less invasive and allows faster recovery than surgery, but its applicability depends on disease stage [21]. Current management of endoscopically diagnosed “atrophic metaplasia” relies primarily on routine follow-up and lacks effective risk stratification tools. This one-size-fits-all approach risks missed intervention opportunities for high-risk patients [22]. CIN testing offers a new pathway for early-stage risk assessment: CIN-positive patients may benefit from intensified, personalized monitoring, while CIN-negative individuals could safely undergo extended surveillance intervals. Such stratification would optimize resource allocation and focus attention on truly high-risk populations.

Longitudinal data from our cohort further highlight CIN’s role in predicting postoperative recurrence. Among 84 patients who underwent endoscopic submucosal dissection (ESD), CIN-positive cases had a 19.57-fold higher risk of metachronous or heterotopic high-risk lesions (p = 0.0039). Only one of the 18 recurrent cases was CIN-negative, likely due to sampling bias, and CIN achieved an AUC of 0.80 for predicting recurrence. These results suggest that CIN is not only a predictor of precancerous progression but also a strong indicator of postoperative recurrence, reinforcing its role as a persistent molecular driver throughout GC progression.

Another notable finding is that some CIN-positive biopsy sites did not develop cancer. This suggests that CIN may reflect a molecular “field effect” beyond localized lesions, indicating broader genomic instability across the gastric mucosa. This parallels blood-based biomarker mechanisms, where systemic or organ-level signals can indicate susceptibility even when the signal source and eventual cancer site do not fully overlap [18, 19]. If the gastric mucosal microenvironment is likened to “carcinogenic soil,” CIN positivity may mark the entire “field” as predisposed to malignant transformation. The spatial consistency of mutational profiles further supports this hypothesis. Moreover, in patients with postoperative pathological upstaging, preoperative CIN positivity reached 90.91%, including cases where IM was later confirmed as cancer. This underscores CIN’s ability to reveal molecular abnormalities that may be missed by pathology, thereby providing more reliable evidence for clinical decision-making. Collectively, our findings support CIN as a biomarker spanning the entire continuum of GC progression.

The strengths of this study include: (1) Long-term cohort data—although involving 117 patients, intensive sampling over a decade (1,199 sequenced samples) enabled construction of a comprehensive molecular timeline of GC natural history, providing robust evidence that CIN precedes GC and has predictive value; (2) Recurrence prediction—prognostic analysis demonstrated CIN’s ability to predict recurrence risk after ESD, confirming its role across initiation, progression, and recurrence; (3) Clinical translatability—CIN assessment advances prevention to the precancerous stage, facilitating a shift from passive screening to proactive management of high-risk groups in line with precision medicine; (4) Molecular characterization—large-scale sequencing revealed extensive chromosomal alterations at the IM stage, with mutation hotspots on chromosomes 7, 8, 13, and 20, resembling those in dysplasia and GC. These instabilities provide insights into the molecular mechanisms of gastric carcinogenesis.

This study also has some limitations. First, its retrospective design: While our data strongly support CIN as a predictive marker across GC progression, large-scale prospective studies in asymptomatic populations are needed to validate its clinical utility. Second, single-center bias:

Although the study prioritized depth of longitudinal sampling (n = 1,119 samples), the limited number of patients (n = 117) and single-center design may introduce selection bias, necessitating multicenter validation. Third, technical variability: CIN detection relied on archived tissue blocks, whose quality may be influenced by fixation, embedding, storage, and sampling depth, potentially introducing variability. Finally, sample failure: Despite strict quality control, some samples failed sequencing, possibly due to block age or sampling site.

## Contributors

Shao-wei Li, Xin-yu Fu, Can Dai, Hao Liu, Jia-xiu Ying, Jin-bang Peng, Jia-cheng Li, Li-na Fang, Xian-bin Zhou, Ya-qi Song, Shi-wen Xu, Shen-ping Tang, Ling-ling Yan, Yuhan Lu, Bin-bin Gu, Liang-min Zhang, Shan-jing Xu, Xia Chen, Ya-hong Chen, Yu Zhang, Li-ping Ye, Mei-fu Gan, and Xin-li Mao developed the protocol, conducted the review, and revised the final draft of the manuscript. All authors collected data, provided input for the protocol, and revised the draft of the manuscript, approving the final version. Li-ping Ye, Shao-wei Li, and Xin-li Mao are the guarantors.

## Ethics

The written patients informed consent was waived by the Ethics Review Committee of Taizhou Hospital, Zhejiang Province (No. K20201205). This manuscript did not involve animal experiences or interventions. Patient and public involvement Patients and/or the public were not involved in the design, or conduct, or reporting, or dissemination plans of this research.

## Data availability

Data are available on reasonable request. All data analysed are available in proper databases depending on publisher.

## Abbreviations

AUC: Area Under the Curve
CBS: Circular Binary Segmentation
CI: Confidence Interval
CIN: Chromosomal Instability
CNA: Copy Number Alteration
CNVs: Copy Number Variations
EGC: Early Gastric Cancer
ESD: Endoscopic Submucosal Dissection
FFPE: Formalin-Fixed Paraffin-Embedded
GC: Gastric Cancer
H&E: Hematoxylin and Eosin
HR: Hazard Ratio
HGIN: High-grade Intraepithelial Neoplasia
IM: Intestinal Metaplasia
LC-WGS: Low-coverage Whole-genome Sequencing
LGIN: Low-grade Intraepithelial Neoplasia
MAD: Median Absolute Deviation
WGS: Whole-genome Sequencing

## Data Availability

All data produced in the present study are available upon reasonable request to the authors.

