## Supplementary materials for "Longitudinal Tracking of Chromosomal Instability Informs Timely Intervention in the Gastric Precancerous Cascade"

### SUPPLEMENTARY MATERIALS AND METHODS

#### DNA Extraction

Genomic DNA was extracted from formalin-fixed, paraffin-embedded (FFPE) tissue samples using the QIAamp DNA Mini Kit (Qiagen, Courtaboeuf, France) according to the manufacturer's instructions, ensuring standardized processing and consistency across samples. This method is widely used for its efficiency and reliability in obtaining high-quality DNA suitable for downstream applications. DNA concentration and purity were measured using a NanoView instrument (GE Healthcare, Orsay, France) with Nanodrop spectrophotometry, enabling accurate quantification and assessment of DNA integrity. DNA quality was further validated by 0.8% agarose gel electrophoresis, which separates DNA fragments under an electric field based on size and charge.

#### Low-Coverage Whole Genome Sequencing

Low-coverage whole genome sequencing (LC-WGS) was performed using libraries constructed with the Kapa Hyper Prep Kit (Roche, CA, USA) and custom adapters (Integrated DNA Technologies, CA, USA). Input DNA ranged from 50 to 800 ng (median: 401 ng). Libraries were pooled in batches of 22 for cost-efficient low-pass sequencing. Pooled libraries were sequenced on an Illumina HiSeq X10 platform (Illumina, USA) using 150 bp paired-end reads in a single lane to maximize throughput and efficiency. Copy number alterations were inferred using the Ultrasensitive Chromosomal Aneuploidy Detector pipeline, which detects genome-wide copy number variations (CNVs) by analyzing log ratios of relative copy numbers between adjacent genomic bins. Samples with a median absolute deviation (MAD) > 0.38 were considered low quality and excluded.

**Copy Number Analysis and Chromosomal Instability (CIN) Score Calculation**

Genomic breakpoints and copy number changes were identified using the Circular Binary Segmentation (CBS) algorithm implemented in the R package DNACopy. Each sample was sequenced to a minimum of 10 million paired-end reads and aligned to the human reference genome (hg19) using the BWA algorithm. Genome-wide coverage depth was assessed with the *mpileup* function in SAMtools, and average coverage was calculated within non-overlapping 200 kb bins for comprehensive evaluation of copy number changes. Coverage values were normalized across datasets, and Z-scores were computed for each bin to quantify deviations from the mean. A Z-score > 3 was defined as amplification (copy number gain), and a Z-score < –3 as deletion (copy number loss). This threshold-based approach provided a practical and interpretable framework for identifying copy number alterations. The Z-score for each bin was derived by comparing its normalized coverage with the overall distribution across samples, representing the difference between observed and mean coverage, normalized by the standard deviation of coverage values (see formula below), where $V_{tumor}$ and $V_{control}$ denote coverage values in the test and reference samples, respectively.

$$Z = \frac{V_{tumor} - average(V_{control})}{stdev(V_{control})}$$

**Clonal Evolution**

Phylogenetic analysis was performed to trace the clonal evolution of CIN. Based on chromosomal aberration data from the Ultrasensitive Chromosomal Aneuploidy Detector, chromosomal arm amplifications were coded as "1," deletions as "–1," and no change as "0." Analyses were conducted in R using the *phangorn* and *ape* packages. Genetic distances between samples were calculated with the Hamming distance, and an initial phylogenetic tree was constructed using the neighbor-joining method. Refinement was carried out under the maximum parsimony criterion with the *Pratchet* function (maximum 1000 iterations), followed by branch-length optimization. Nodal support was assessed with 1000 bootstrap replicates.

**Statistical Analysis**

Statistical analyses were conducted in R (version 4.1.1). Normally distributed continuous variables are expressed as mean ± standard deviation (SD), whereas non-normally distributed variables are presented as median (interquartile range [IQR]). Categorical variables are summarized as frequency and percentage [n (%)]. One-way analysis of variance (ANOVA) was used to compare normally distributed continuous variables across groups, with post hoc pairwise comparisons performed using the Newman–Keuls method. Fisher's exact test was applied to evaluate associations between categorical variables. Disease-free survival (DFS) was analyzed with Kaplan–Meier curves and compared using the log-rank test. Cox proportional hazards regression was used to estimate hazard ratios (HRs). A two-sided p-value $< 0.05$ was considered statistically significant.

**Supplementary Materials and Methods References**

1. Li, Heng, and Richard Durbin. Fast and accurate long-read alignment with Burrows-Wheeler transform. Bioinformatics (Oxford, England) vol. 26,5 (2010): 589-95.

2. Li, H., Handsaker, B., Wysoker, A., Fennell, T., Ruan, J., Homer, N., Marth, G., Abecasis, G., Durbin, R., & 1000 Genome Project Data Processing Subgroup (2009).The Sequence Alignment/Map format and SAMtools. Bioinformatics, 25(16), 2078–2079

3. Wilgenbusch JC, Swofford D. Inferring evolutionary trees with PAUP*. Curr Protoc Bioinformatics 2003;Chapter 6:Unit 6 4.

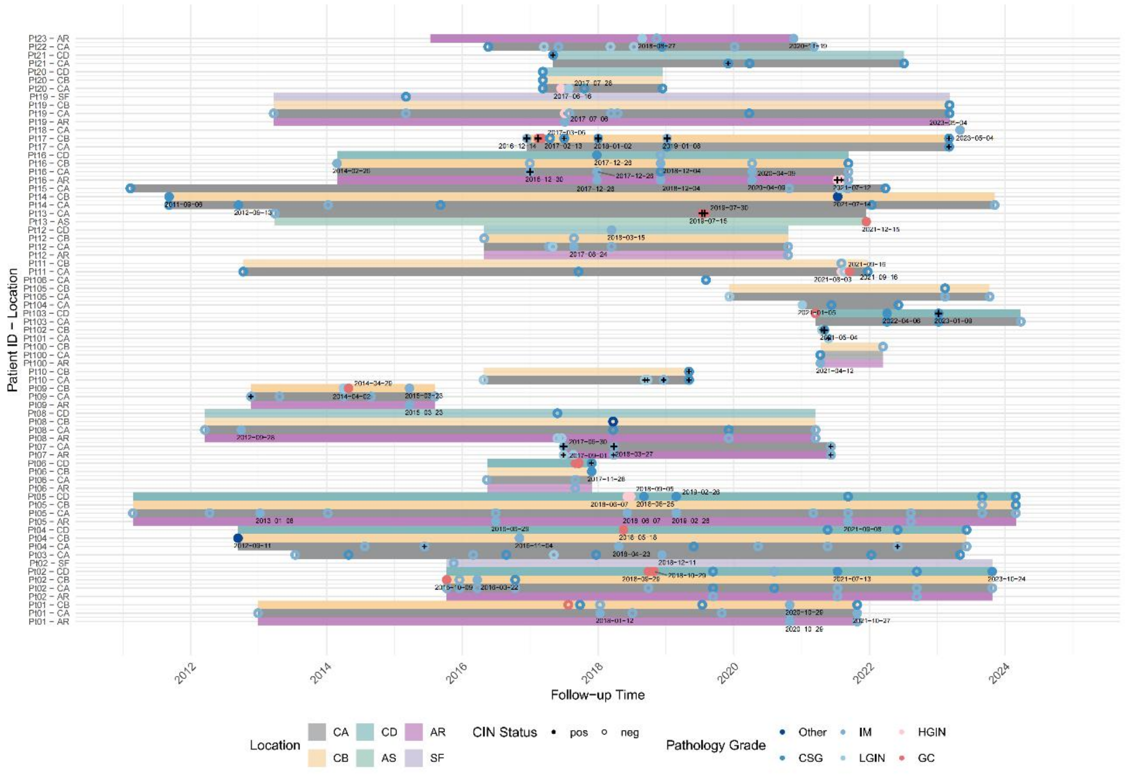


**Figure S1.** Sample Time Lane Chart 1.

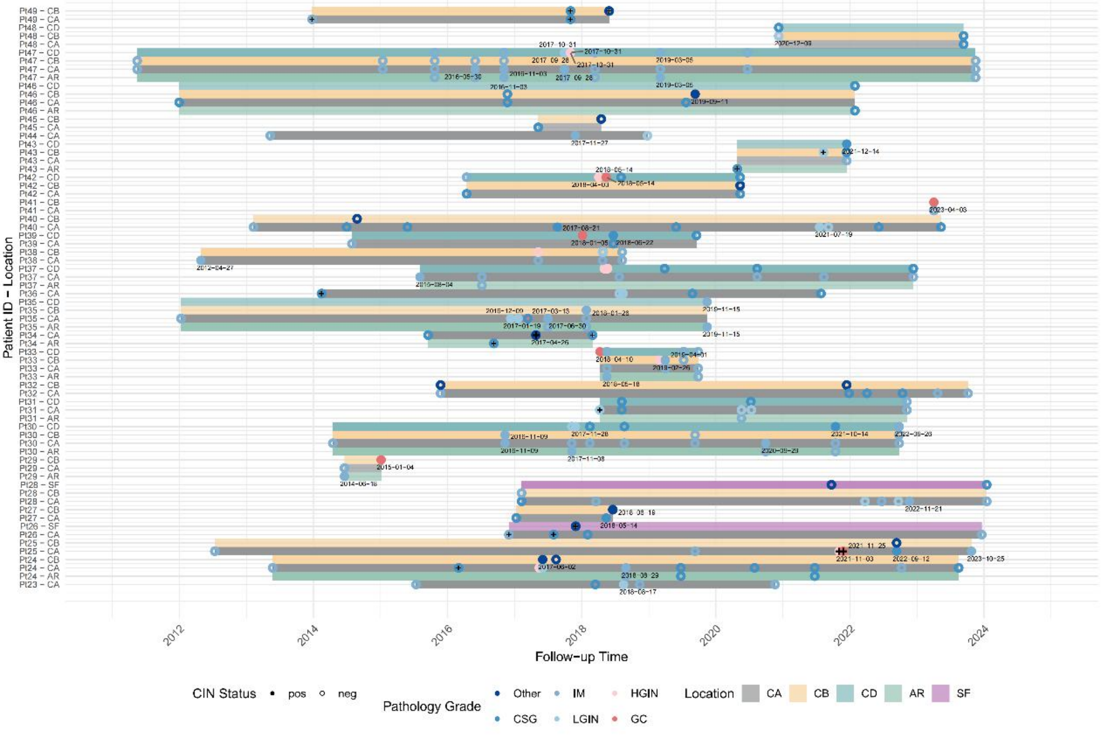


**Figure S2.** Sample Time Lane Chart 2.

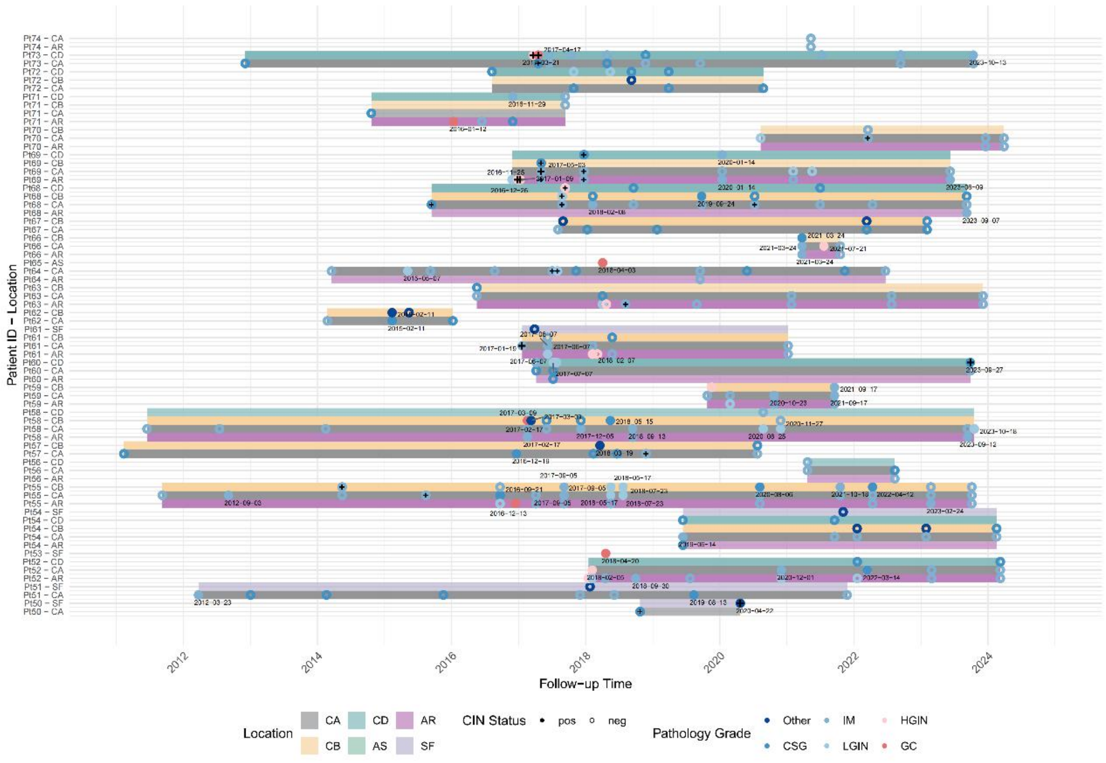


**Figure S3.** Sample Time Lane Chart 3.

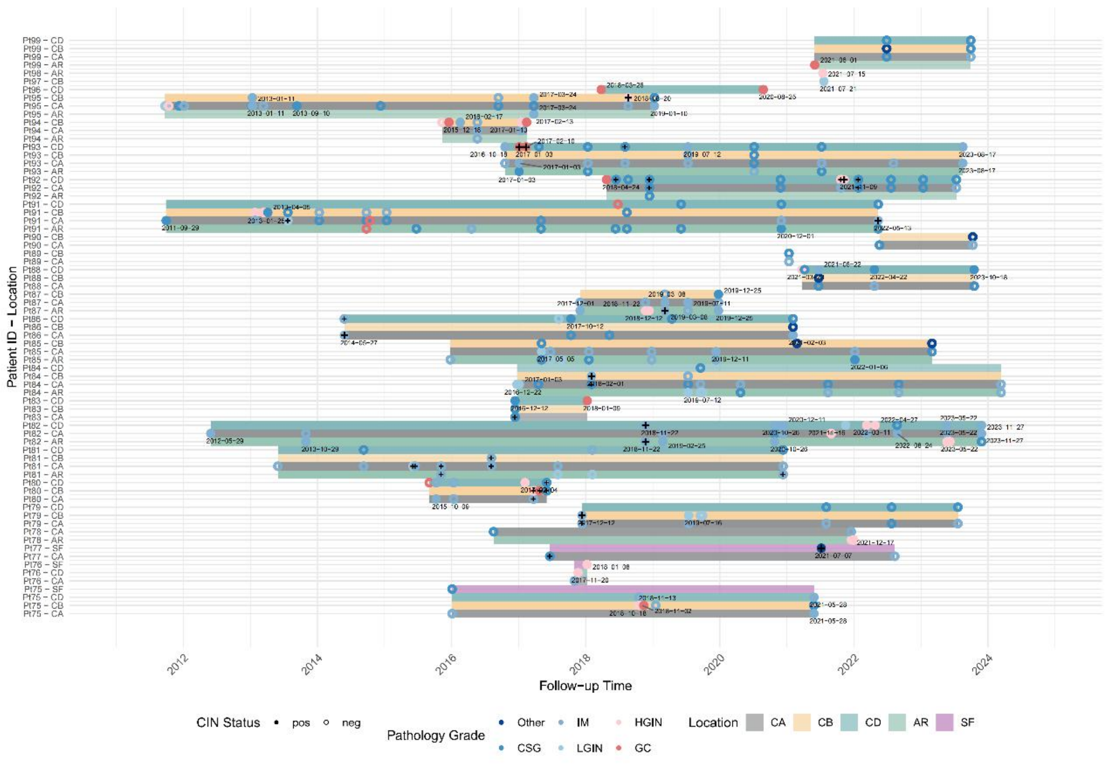


**Figure S4.** Sample Time Lane Chart 4. CA: Gastric antrum; CD: Cardia; AR: Gastric angle; CB: Gastric body; SF: Gastric fundus; AS: Anastomosis; Other(Polyps and stromal tumors);

IM(Intestinal metaplasia);CSG(chronic gastritis);HGIN(High-grade Intraepithelial Neoplasia);LGIN(Low-grade Intraepithelial Neoplasia);GC(gastric cancer)

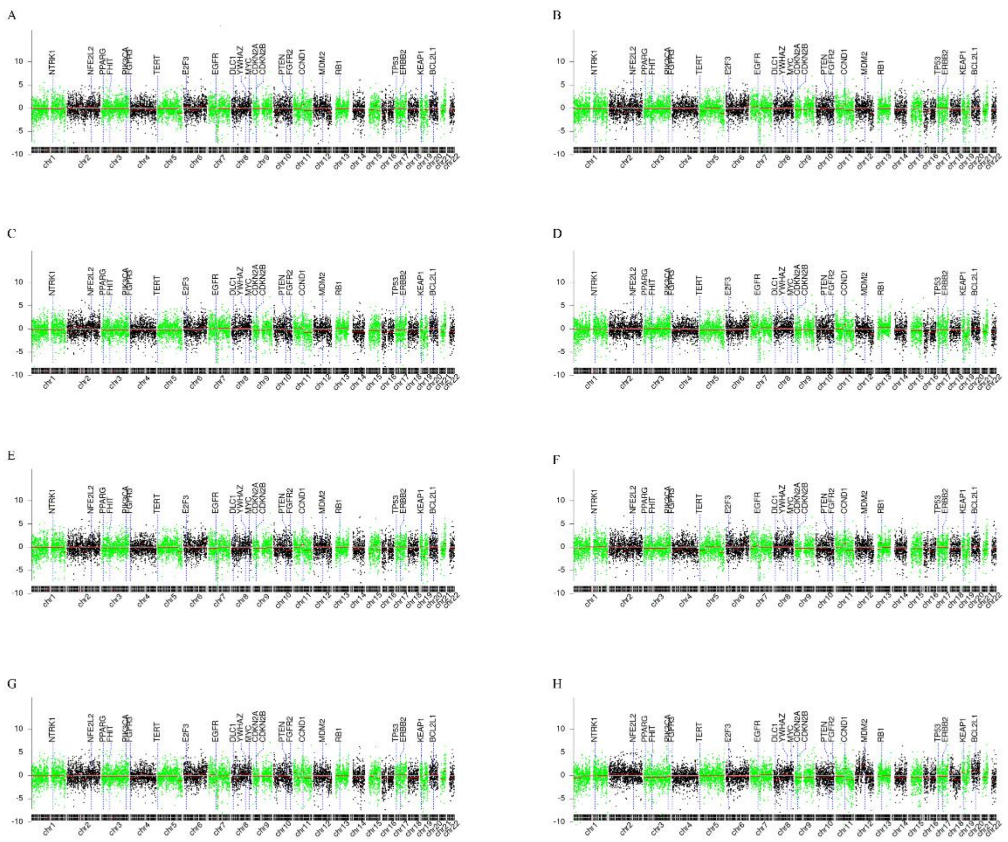


**Figure S5.** Chromosome based display of local gene mutations. A-H: Analysis of mutated genes at chromosomal loci in some samples.

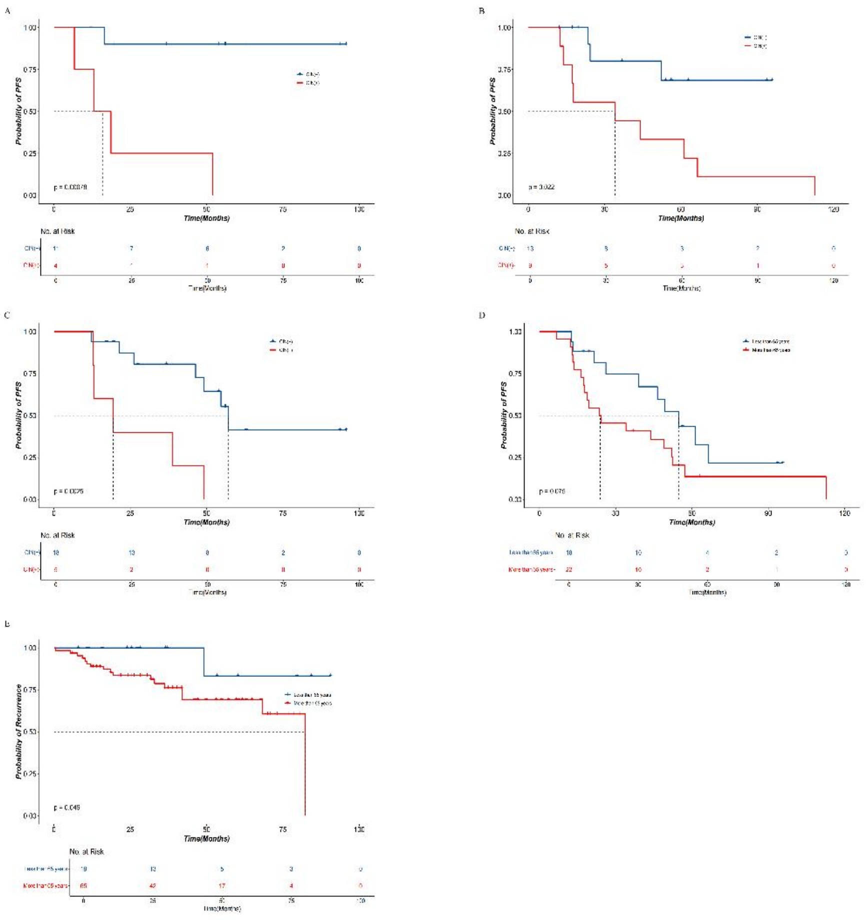


**Figure S6.** KM curve chart. A. KM curve of progression to gastric cancer; B.KM curve of progression to High-grade Intraepithelial Neoplasia; C. KM curve of progression to Low-grade Intraepithelial Neoplasia; D. KM Curve by Age Groups in the Progression Cohort; E. KM Curve by Age Groups in the Recurrence Cohort.

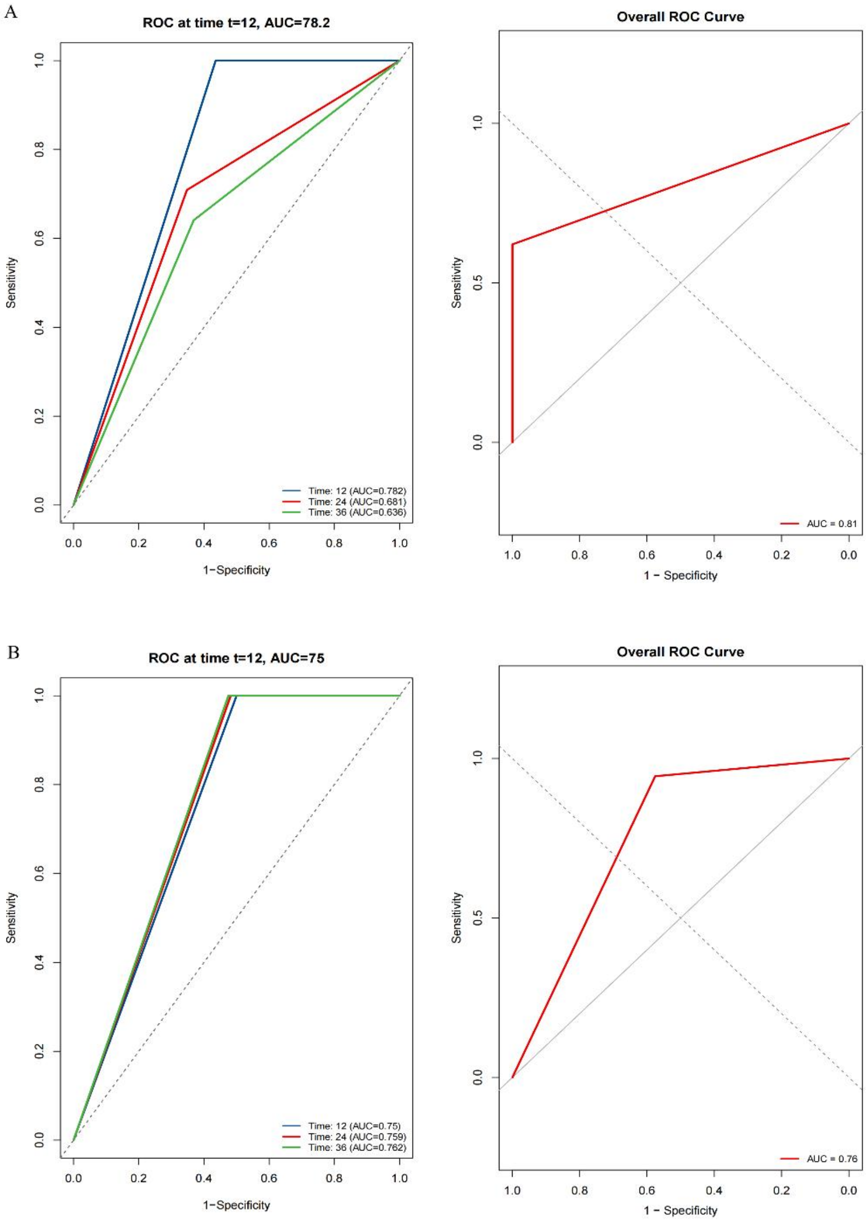


**Figure S7.** ROC Plot for the Predictive Performance of the CIN Marker in the Progression Cohort and Recurrence Cohort. A. ROC Plot for the Predictive Performance of the CIN Marker in the Progression Cohort. B. ROC Plot for the Predictive Performance of the CIN Marker in the Recurrence Cohort.

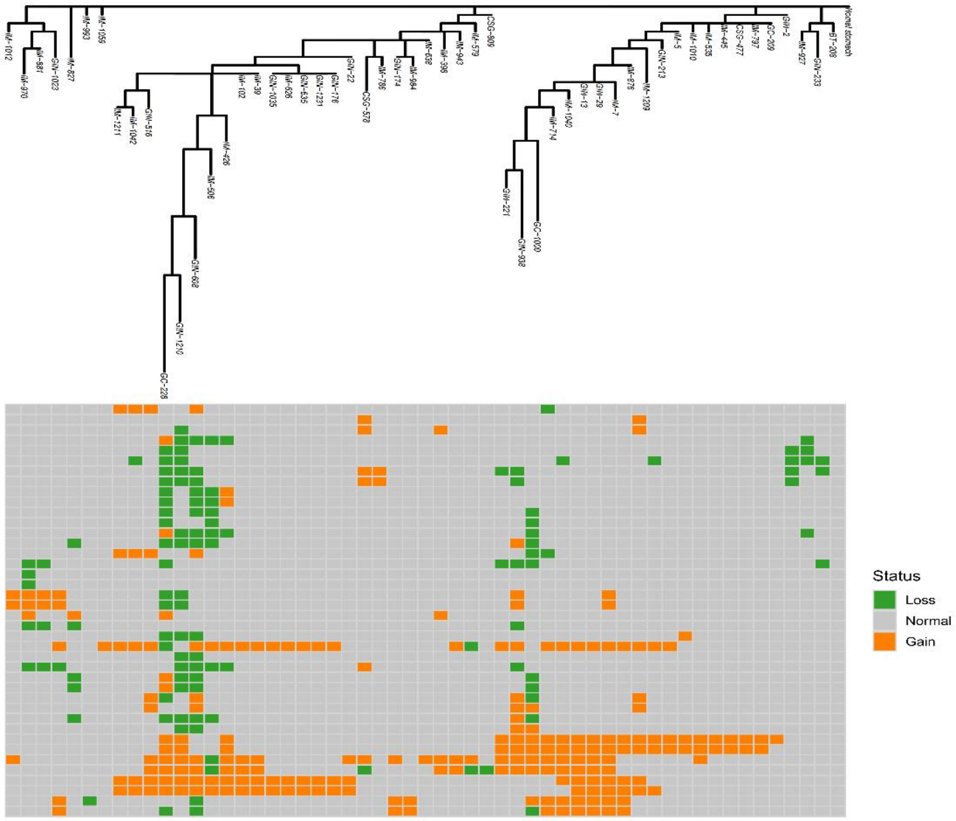


**Figure S8.** Clonal Evolution Analysis of Samples with Pathological Recurrence.

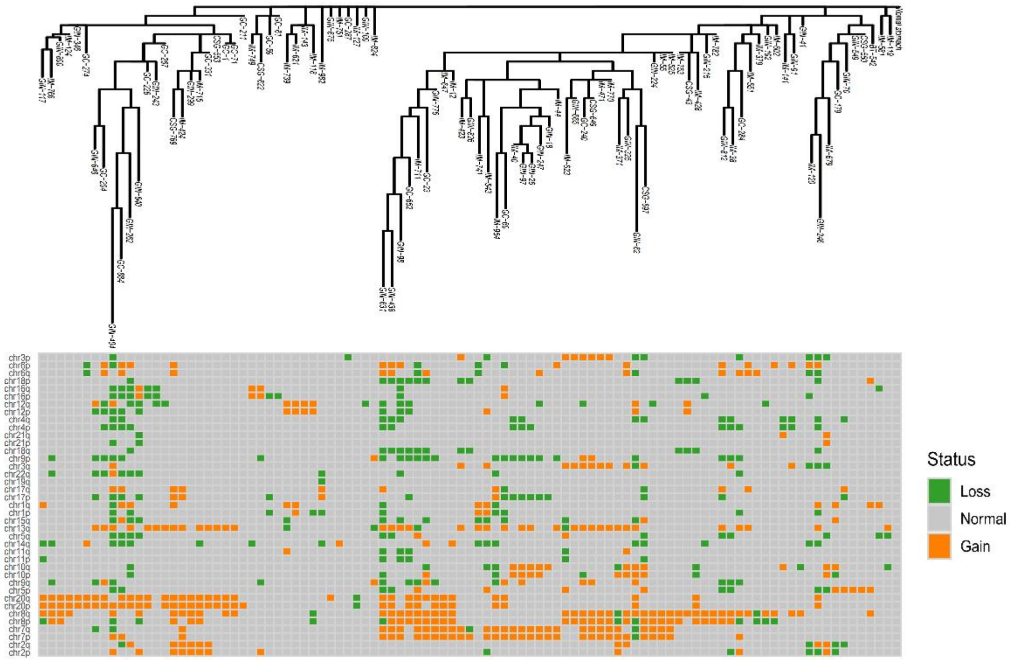


**Figure S9.** Clonal Evolution Analysis of Samples with Pathological Progression.

**Table S1.** Univariate Analysis on the Progression and Recurrence of Gastric Cancer

| | **Progression** | | **Recurrence** | |
|---|---|---|---|---|
| | **HR (95% CI)** | ***P*** | **HR (95% CI)** | ***P*** |
| CIN (+) | 2.55 (1.19-5.47) | 0.016 | 19.57 (2.59-147.60) | 0.004 |
| Age > 65 | 2.01 (0.92-4.38) | 0.081 | 6.40 (0.82-50.03) | 0.077 |
| Female | 0.81 (0.37-1.80) | 0.607 | — | — |
| HP (+) | 0.81 (0.24-2.73) | 0.730 | 1.72 (0.66-4.50) | 0.267 |
